# Predicting Functional Changes in Down Syndrome During the COVID-19 Pandemic: The Role of Biopsychosocial Determinants of Health

**DOI:** 10.64898/2026.05.19.26353577

**Authors:** Ester Jo, Carla Wall, Lindsay K. Allen, Naomi Wheeler, Nicole Baumer, Allison D’Aguilar, Timothy P. York, George Capone, Colleen Jackson-Cook, Ananda B. Amstadter, Ruth C. Brown

## Abstract

**Background:** Biopsychosocial factors associated with functional changes, including changes in personality, communication, movement, and weight, were evaluated in individuals with Down syndrome (DS) during the COVID-19 pandemic.

**Method:** Caregivers of individuals with DS (aged ≥12, n = 118) completed an online survey. Elastic net regression with bootstrap resampling assessed 31 candidate predictors.

**Results:** Pandemic-related mental health was most strongly associated with functional changes (β = 0.388). Healthcare access barriers were also reliably selected: inability to access mental health treatment, difficulty affording insurance, difficulty accessing specialists, and residing in a low-income health professional shortage area. The model explained 35.2% of variance.

**Conclusions:** Mental health and healthcare access barriers were biopsychosocial correlates of functional changes for people with DS during COVID-19.

---

Down Syndrome is a genetic disorder characterized by three copies of chromosome 21 and occurs in approximately 1 in every 600 - 700 live births (Mai et al., 2019). Down syndrome is associated with a wide range of medical, cognitive, social, and functional phenotypes including, intellectual disability (ID), early onset Alzheimer’s disease, immune dysfunction, thyroid dysfunction, and sleep apnea (Carfì et al., 2020). People with Down syndrome also exhibit a wide range of functional skills and abilities across their lifespan (de Graaf et al., 2019; Matthews et al., 2018). Functional changes, including changes in personality, communication, motor movement, and weight have been found to be associated with other health outcomes (Matthews et al., 2018), and may be important observable markers for the emergence of underlying medical or mental health conditions (Forster-Gibson, 2019).

People with Down syndrome also face many increased socially-determined barriers to social and physical health. People with Down syndrome are often described as being particularly socially motivated (Schworer et al., 2021) and yet they tend to experience limited opportunities for social inclusion and engagement (Guralnick et al., 2009), increasing risk for mental health outcomes associated with loneliness. Before the pandemic, studies found that caregivers of patients with Down syndrome reported difficulties navigating complex healthcare systems and reported challenges in finding mental health specialists who were both knowledgeable about Down syndrome and accessible within a reasonable distance (King et al., 2022). These barriers can contribute to worsened health outcomes, including higher rates of anxiety, depression, and social isolation (McGrath et al., 2011).

The COVID-19 pandemic had a profound impact on the global population, leading to significant shifts in daily lifestyle and contributing to a wide array of mental and physical health changes. Disruptions to daily routines were associated with heightened domestic violence, drug abuse, decreased physical activity, unhealthy eating habits, and more sedentary lifestyles (Grasso et al., 2021). However, individuals with disabilities, especially those with Down syndrome, faced unique challenges, including the reduction of critical disability programs they rely on due to COVID-19 containment measures (Hartley et al., 2022; Villani et al., 2020), increased vulnerability to severe health consequences (Espinosa, 2020), and threats of medical rationing based on disability (Bagenstos, 2020). Additionally, some adults with Down Syndrome encountered challenges due to inaccessible technology that failed to meet their communication, sensory, and cognitive needs, thereby impacting their ability to fully benefit from the rapid shifts to telehealth or virtual services (Annaswamy et al., 2020; Hartley et al., 2022).

Unfortunately, only a few investigators have studied the impact of the COVID-19 pandemic on individuals with Down Syndrome (Hartley et al., 2022; Rubenstein et al., 2023; Sideropoulos et al., 2023; Villani et al., 2020). Studies have demonstrated that changes in employment or day programs due to the pandemic were a significant source of stress for many. For instance, 51% of adults with Down Syndrome reported these changes as ‘slightly’ to ‘moderately’ stressful, and 21% found them ‘very’ to ‘extremely’ stressful (Hartley et al, 2022). Bond et al. (2020) further emphasize that older adults with intellectual disabilities, particularly those with Down syndrome, are more likely to experience anxiety and depressive symptoms, which are closely associated with modifiable biopsychosocial factors such as loneliness, sleep difficulties, and aggressive challenging behavior. Prior to the COVID-19 pandemic, studies have shown that adults with Down syndrome commonly experience mood and behavior problems, including depression and anxiety symptoms (Mantry et al., 2007; Tassé et al., 2016).

Documenting and understanding the impacts of the COVID-19 pandemic on underserved populations and those with preexisting vulnerabilities is essential for improving the response in the event of future widespread public health emergencies (Thomas et al., 2022). The relative lack of studies on the impact of the pandemic on people with Down syndrome limits our understanding and ability to adequately support people with Down syndrome experiencing long-term effects of the pandemic. This study aims to fill this gap by examining the risk factors associated with functional changes in individuals with Down syndrome. Given the significant changes in daily life and the potential for increased stress and anxiety caused by the pandemic, it is crucial to understand how these biopsychosocial factors might contribute to changes in well-being in this population as measured by observable changes in functional changes. We hypothesize that social determinants of health, including access to healthcare, socio-economic status, and environmental factors, along with prior mental health conditions, will significantly predict functional changes in individuals with Down syndrome during the COVID-19 pandemic.

## Methods

### Participants

Participant data were drawn from a larger mental health surveillance study of parents and caregivers of children or adults with Down Syndrome at BLINDED FOR PEER REVIEW. Participants completed the online survey on REDCap (Research Electronic Data Capture; Harris et al., 2009) from October 2020 through April 2022, between 6 and 25 months after the declared start of the COVID-19 pandemic in the United States (March 2020), with a median of 13.4 months. Enrollment occurred across three main recruitment efforts using invitations sent through Down syndrome-specific registries and listservs, and the NIH DS-Connect registry (Peprah et al., 2015). Survey timestamps were used to calculate the time since the start of the pandemic and these were binned into one of three enrollment waves. Wave 1 occurred within 10 months of the start of the pandemic, Wave 2 occurred during months 11-20, and Wave 3 occurred during months 21-30 of the pandemic. Inclusion criteria included caregiver age of 18 years or older, English- or Spanish-speaking parents and/or legally authorized primary caregivers of children or adults with Down syndrome aged 12 years and older, and residing in the United States. The Institutional Review Board approved all study procedures at BLINDED FOR PEER REVIEW.

## Measures

### Demographics

Participants completed a study-designed questionnaire to collect basic demographic information, including the age, sex, race, ethnicity, and income of both the caregiver and the child, and caregiver proxy reports of co-occurring health conditions, maladaptive behavior, and posttraumatic stress symptoms.

### Pandemic Impact

Mental health impacts related to the COVID-19 pandemic were assessed using a modified version of the Epidemic-Pandemic Impacts Inventory (EPII; Grasso, Briggs-Gowan, Carter, Goldstein, Ford, 2021). Initially, the EPII provided response options of “Yes (me),” “Yes (person in home),” “No,” and “N/A.” To specifically capture the experience of individuals with Down syndrome, an additional response category, “Yes (person with Down syndrome),” was added. Items evaluated pandemic-related increases in behavioral or emotional problems, sleep difficulties, general mental health symptoms, access difficulties, and satisfaction with mental health treatment changes.

Responses indicating increases in behavioral/emotional problems or mental health symptoms were combined into a single mental health impact variable. Similarly, increased sleep difficulties, nightmares, and general sleep problems were combined into a sleep impact variable.

### Observed Functional Changes

Items from the “Notable Significant Changes Observed by Others” subscale of the National Task Group on Intellectual Disabilities and Dementia Practices – Early Detection Screen for Dementia (NTG-EDSD; Esralew et al., 2013) were administered as a proxy for functional changes. This subscale was selected for brevity and due to studies showing that mood and cognitive symptoms may not be verbalized in people with Down syndrome, often requiring the need for observations of behavioral changes by caregivers (Forster-Gibson, 2019). Items assessed whether the caregiver observed new or worsening difficulties in personality, friendliness, gait, attentiveness, and weight. Each item was rated on a 4-point scale: “This has always been the case for this person and it has not changed since the beginning of the pandemic;” “This has always been the case, but it has gotten worse since the beginning of the pandemic;” “This is a new symptom since the beginning of the pandemic;” and “This symptom does not apply to this person.”

This scoring strategy allows for comparison of symptoms against the person’s background level of functioning (Silverman et al., 2021). In this study functional changes were operationalized as the count of symptoms caregivers endorsed as either “Always but worse” or “New symptom since the beginning of the pandemic,” yielding a 0–5 score representing the number of domains in which deterioration was observed.

### Data Analysis Plan

A set of 31 predictors was selected based on theoretical relevance (e.g., Fong & Iarocci, 2020; Moynihan et al., 2021; Samji et al., 2022), including demographic factors (e.g., pandemic wave timing, child age), socioeconomic indicators (e.g., low-income area with health professional shortages, difficulty paying for health insurance), mental health diagnoses (e.g., psychosis, regression or disintegrative disorder), and pandemic-related disruptions (e.g., unable to access mental health treatment since the pandemic, pandemic-related mental health impacts, pandemic-related sleep impacts). Composite variables included pandemic-related mental health impacts (combined behavioral/emotional and mental health symptoms). Given the broad range and interrelatedness of potential predictors, traditional regression methods may inadequately address multicollinearity and the complexity inherent in these data. To assess multicollinearity among predictors, we computed Variance Inflation Factors (VIFs) using a standard linear model. The mean VIF across all predictors was 2.00, with a maximum value of 3.83. No predictors exceeded the commonly accepted thresholds of 5 or 10, indicating no substantial multicollinearity in the dataset. However, given the large number of predictors and the moderate intercorrelations, we employed Elastic Net regression to guard against overfitting and to perform variable selection. The Elastic Net approach, which combines L1 and L2 regularization, is particularly well-suited for models with potentially correlated predictors and enhances the model’s generalizability.

Missing data were imputed using the ‘micè package in R. The multiple imputation methods were tailored to variable type: predictive mean matching (using ‘pmm’) for continuous and binary variables, proportional odds regression (using ‘polr’) for ordered categorical variables, multinomial logistic regression (using ‘polyreg’) for nominal categorical variables, and ‘midastouch’ for rare binary variables.

The analysis was conducted in R (version 4.5.1) using the tidymodels (version 1.3.0) and glmnet (version4.1-10) packages. Data were preprocessed by removing zero-variance predictors, normalizing numeric predictors, and creating dummy variables for categorical predictors. Only additive (main effect) contributions were modeled. A 3-fold cross-validation, stratified by the outcome variable, was used to tune the elastic net hyperparameters and to estimate expected model performance. Hyperparameters were tuned over a regular grid search of 125 combinations spanning 25 penalty values and 5 mixture values. The best-performing model was selected using the minimum cross-validated RMSE criterion. The optimal model was selected based on the minimum cross-validated root mean squared error (RMSE), yielding λ = 0.215 and α = 0.550. This final model was then fit to the complete dataset to obtain predictor coefficients. Model performance was evaluated on the complete dataset using RMSE, R-squared (R²), and mean absolute error (MAE). Non-zero coefficients from the final model were extracted to identify significant predictors.

Because Elastic Net regression does not produce standard errors or p-values for individual coefficients, we used a nonparametric bootstrap procedure to characterize uncertainty and predictor selection stability. One thousand bootstrap resamples of the full dataset were drawn. All 31 predictor coefficients were retained from each iteration, including those shrunk to zero, so that zero-coefficient iterations contributed to the bootstrap averages. For each predictor, we computed the bootstrap-averaged standardized coefficient (β), the 95^th^ percentile confidence interval, and the frequency in which the predictor received a non-zero coefficient. Predictors were considered reliably retained if they had a mean |β| > 0.001 and were selected in at least 50% of bootstrap samples.

## Results

### Demographics Characteristics

The sample (N = 118) was predominantly White (92%), with a mean age of 23 years (SD = 8), and primarily consisted of caregivers who identified as women (87%). Most individuals had a diagnosis of full trisomy 21 (80%).

### Frequency of Functional Changes

Caregivers reported new or worsened functional changes in the person with DS during the pandemic, most commonly related to weight (37%), personality (23%), attentiveness (22%), and friendliness (17%). Changes in gait (6%) and abnormal movement (5%) were less frequently endorsed (see Figure 1). The distribution of observed functional changes reported during the COVID-19 pandemic was positively skewed, with most participants experiencing relatively few changes (Figure 2).

**Figure 1.**
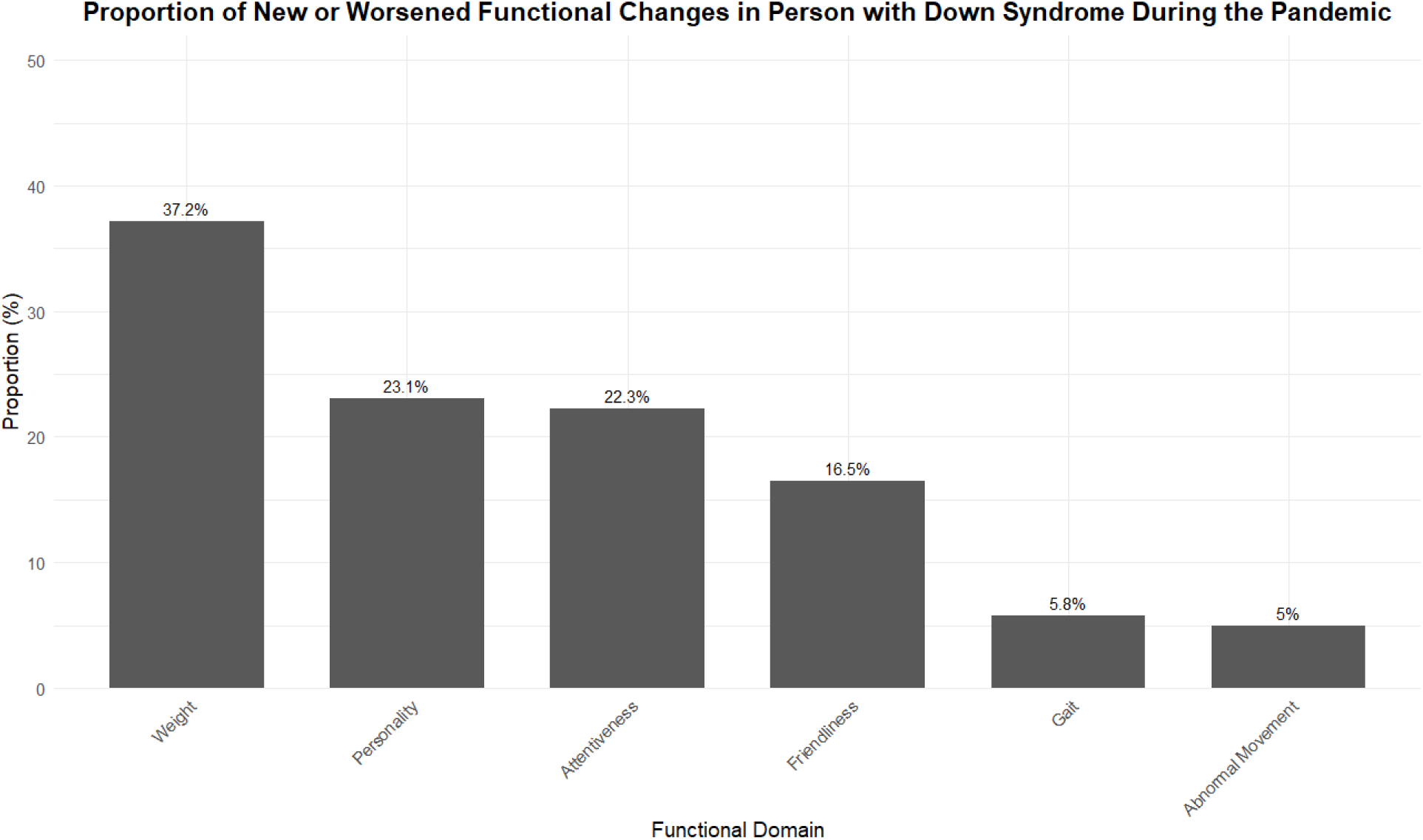
Proportion (%) of Participants Reporting New or Worsened Functional Changes During the COVID-19 Pandemic.

**Figure 2.**
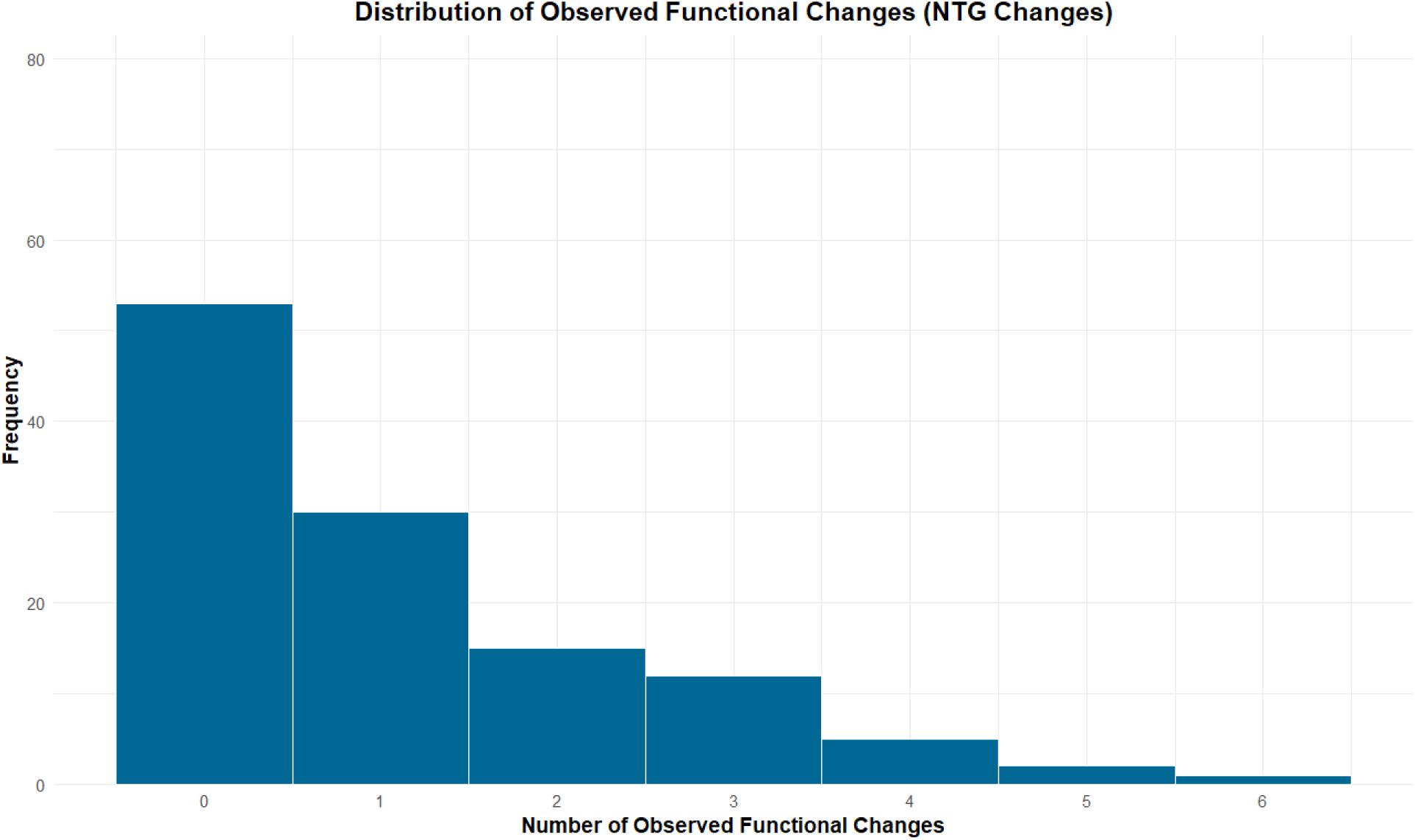
Frequency Distribution of Total Number of Observed Functional Changes Reported During the COVID-19 Pandemic.

Specifically, the largest proportion of participants reported one or fewer functional changes (n = 83, 70.3%). A substantially smaller number of caregivers reported two or more functional changes (n = 35, 29.7%). A small subset of participants reported experiencing four or more changes (n = 8, 6.8%).

### Predictors of Functional Changes

An Elastic Net regression analysis was conducted on the set of 31 theoretically derived predictors of functional changes as measured by the NTG (NTG Changes).

Descriptive statistics of these 31 predictors can be found in Table 1. Of these 31 candidate predictors, 10 were reliably retained, defined as having a bootstrap-averaged |β| > 0.001 and being selected in at least 50% of 1,000 bootstrap resamples (Table 2, Figure 3).

**Table 1.**
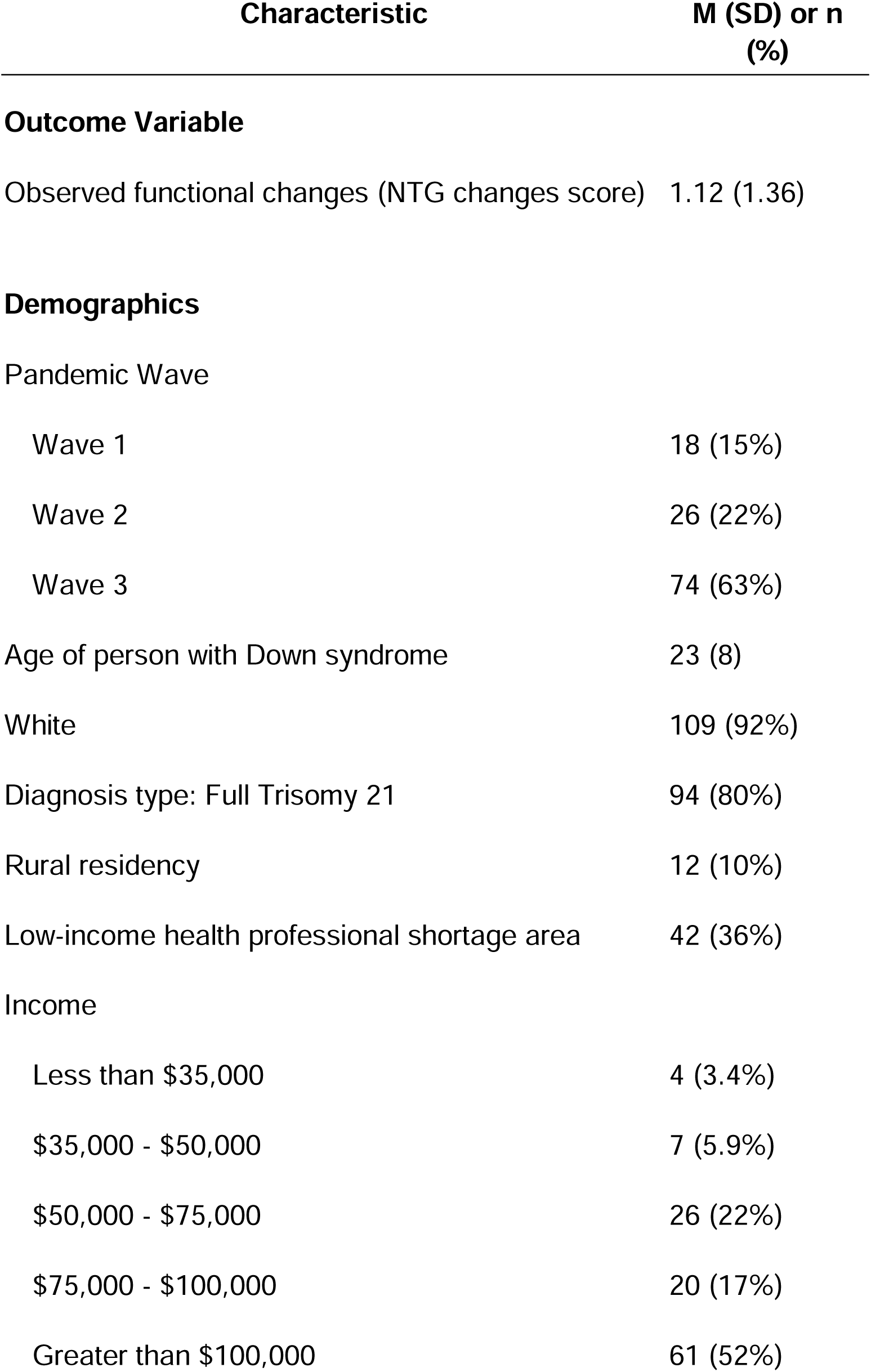
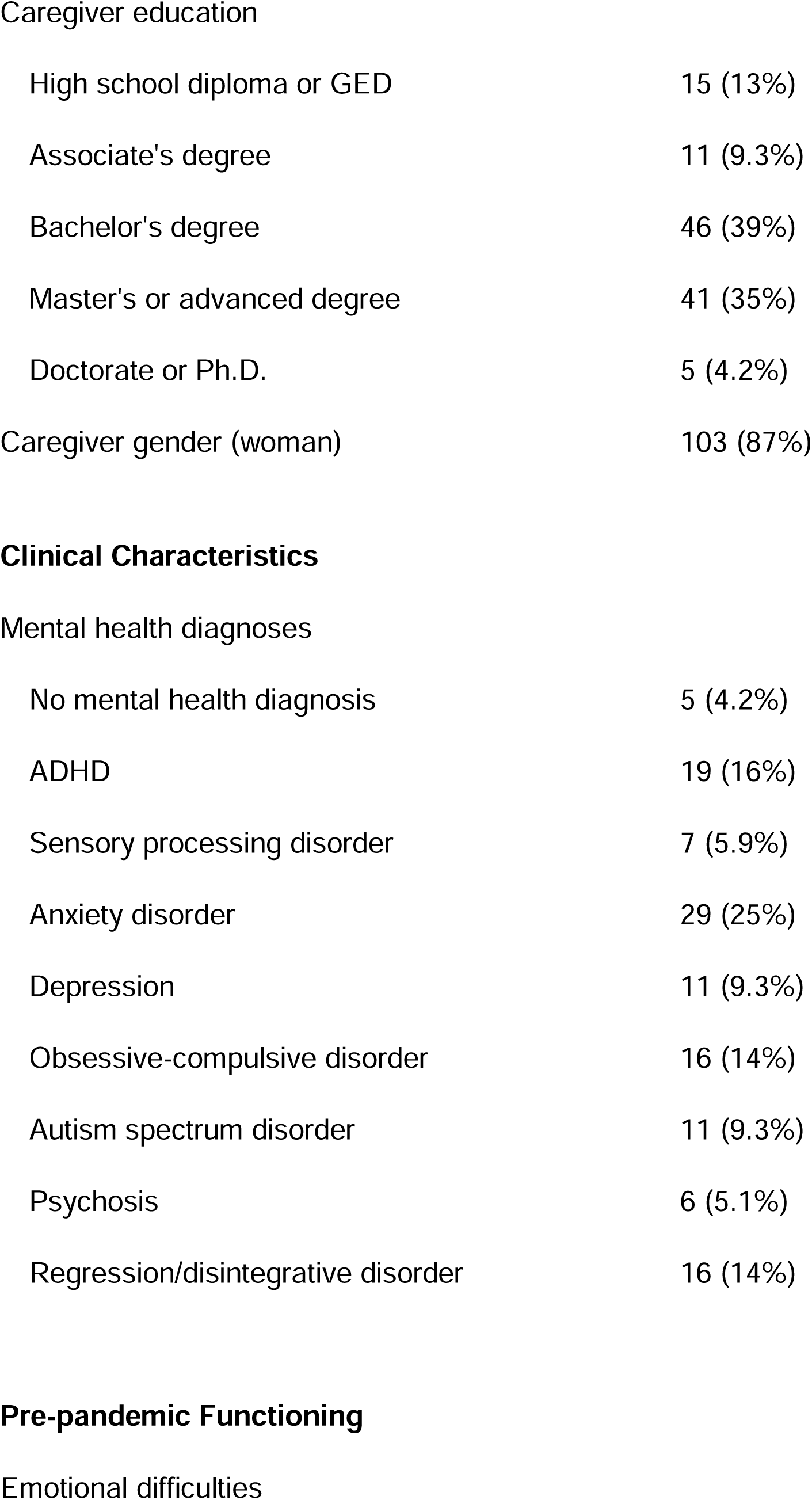
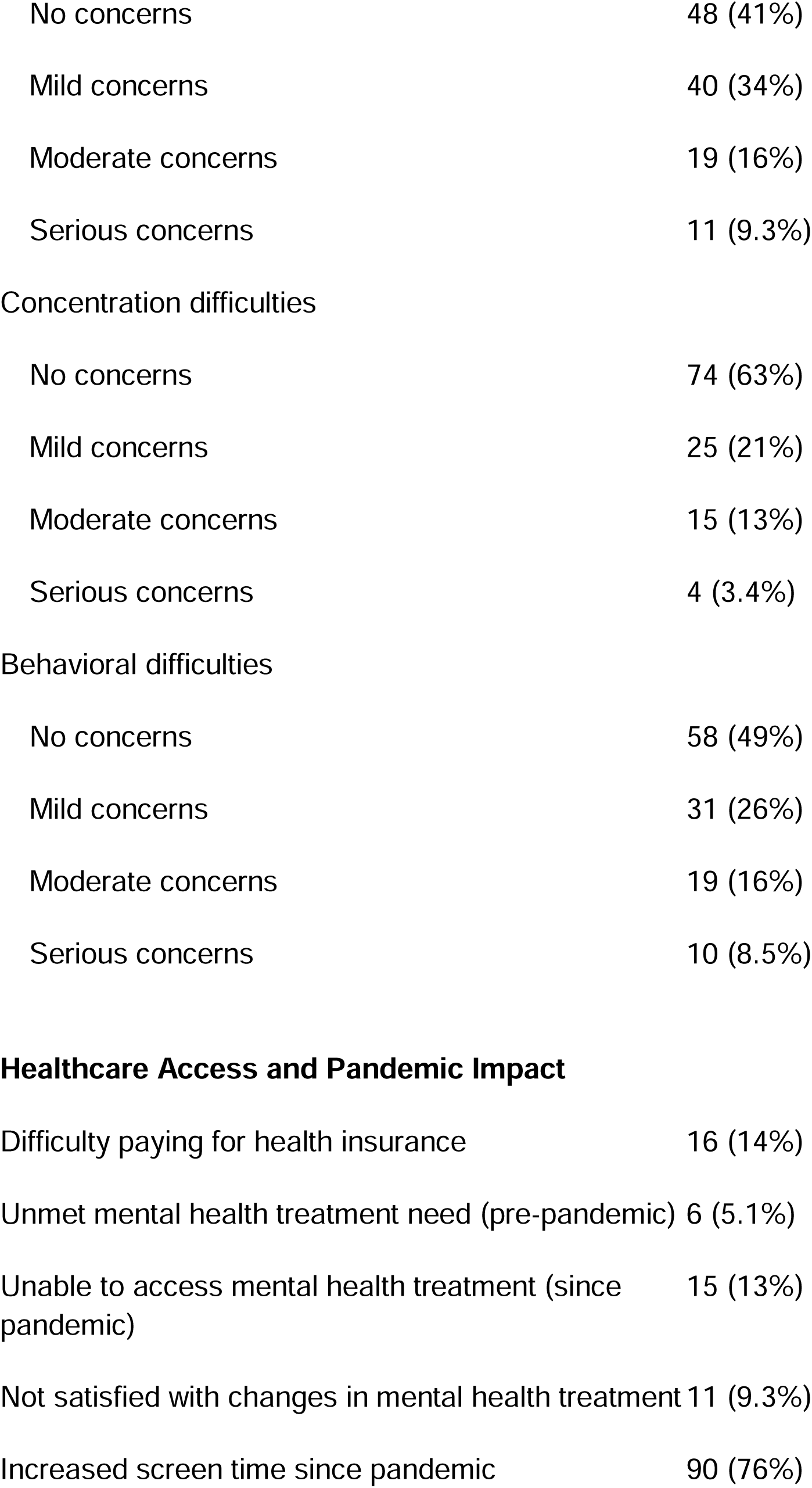
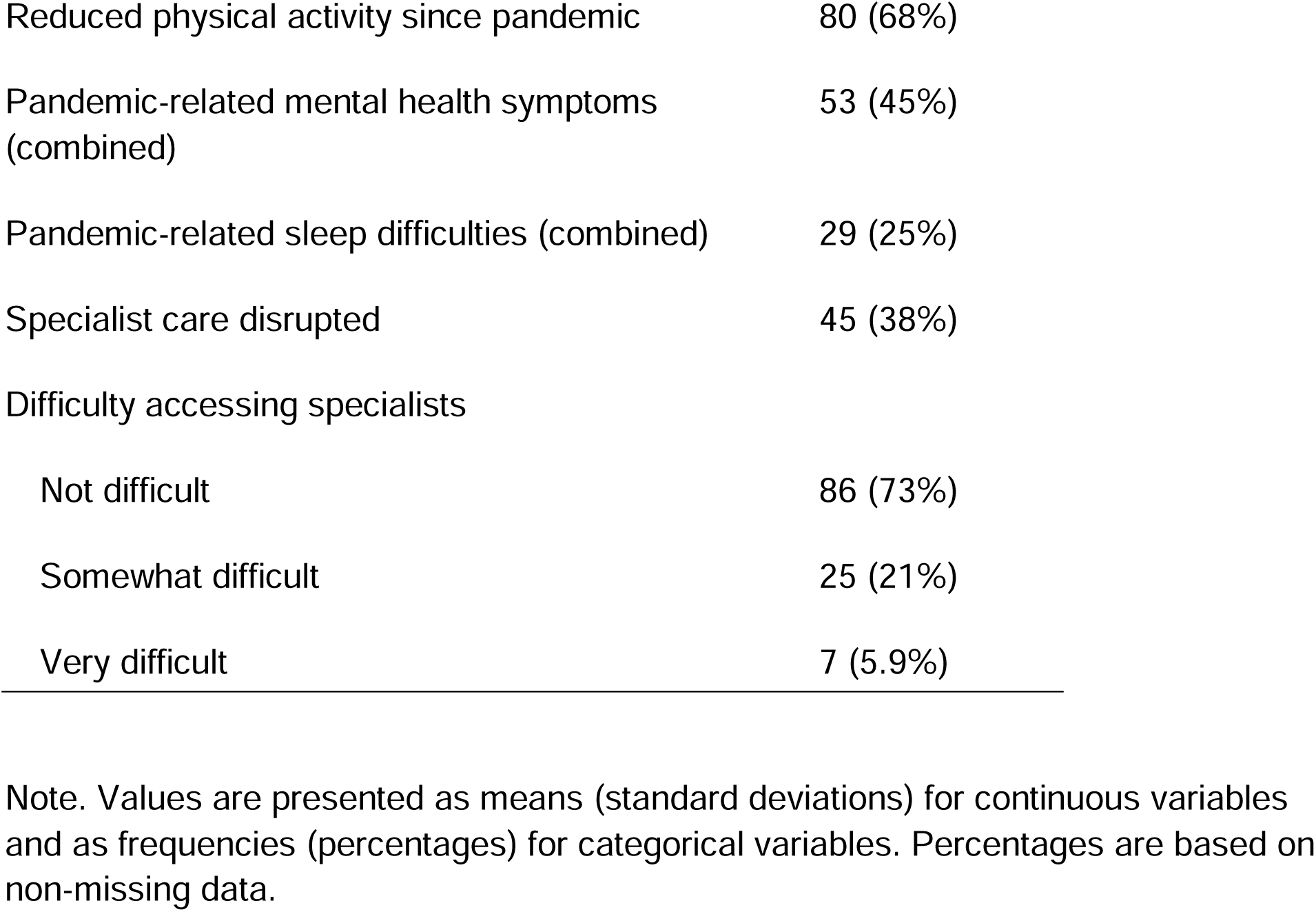
Participant Characteristics (N = 118)

| Characteristic | M (SD) or n (%) |
| --- | --- |
| **Outcome Variable** |  |
| Observed functional changes (NTG changes score) | 1.12 (1.36) |
| **Demographics** |  |
| Pandemic Wave |  |
| Wave 1 | 18 (15%) |
| Wave 2 | 26 (22%) |
| Wave 3 | 74 (63%) |
| Age of person with Down syndrome | 23 (8) |
| White | 109 (92%) |
| Diagnosis type: Full Trisomy 21 | 94 (80%) |
| Rural residency | 12 (10%) |
| Low-income health professional shortage area | 42 (36%) |
| Income |  |
| Less than \$35,000 | 4 (3.4%) |
| \$35,000 - \$50,000 | 7 (5.9%) |
| \$50,000 - \$75,000 | 26 (22%) |
| \$75,000 - \$100,000 | 20 (17%) |
| Greater than \$100,000 | 61 (52%) |
| Caregiver education |  |
| High school diploma or GED | 15 (13%) |
| Associate's degree | 11 (9.3%) |
| Bachelor's degree | 46 (39%) |
| Master's or advanced degree | 41 (35%) |
| Doctorate or Ph.D. | 5 (4.2%) |
| Caregiver gender (woman) | 103 (87%) |
| Mental health diagnoses |  |
| No mental health diagnosis | 5 (4.2%) |
| ADHD | 19 (16%) |
| Sensory processing disorder | 7 (5.9%) |
| Anxiety disorder | 29 (25%) |
| Depression | 11 (9.3%) |
| Obsessive-compulsive disorder | 16 (14%) |
| Autism spectrum disorder | 11 (9.3%) |
| Psychosis | 6 (5.1%) |
| Regression/disintegrative disorder | 16 (14%) |
| Emotional difficulties |  |
| No concerns | 48 (41%) |
| Mild concerns | 40 (34%) |
| Moderate concerns | 19 (16%) |
| Serious concerns | 11 (9.3%) |
| Concentration difficulties |  |
| No concerns | 74 (63%) |
| Mild concerns | 25 (21%) |
| Moderate concerns | 15 (13%) |
| Serious concerns | 4 (3.4%) |
| Behavioral difficulties |  |
| No concerns | 58 (49%) |
| Mild concerns | 31 (26%) |
| Moderate concerns | 19 (16%) |
| Serious concerns | 10 (8.5%) |
| **Healthcare Access and Pandemic Impact** |  |
| Difficulty paying for health insurance | 16 (14%) |
| Unmet mental health treatment need (pre-pandemic) | 6 (5.1%) |
| Unable to access mental health treatment (since pandemic) | 15 (13%) |
| Not satisfied with changes in mental health treatment | 11 (9.3%) |
| Increased screen time since pandemic | 90 (76%) |
| Reduced physical activity since pandemic | 80 (68%) |
| Pandemic-related mental health symptoms (combined) | 53 (45%) |
| Pandemic-related sleep difficulties (combined) | 29 (25%) |
| Specialist care disrupted | 45 (38%) |
| Difficulty accessing specialists |  |
| Not difficult | 86 (73%) |
| Somewhat difficult | 25 (21%) |
| Very difficult | 7 (5.9%) |

Note. Values are presented as means (standard deviations) for continuous variables and as frequencies (percentages) for categorical variables. Percentages are based on non-missing data.

**Table 2.**
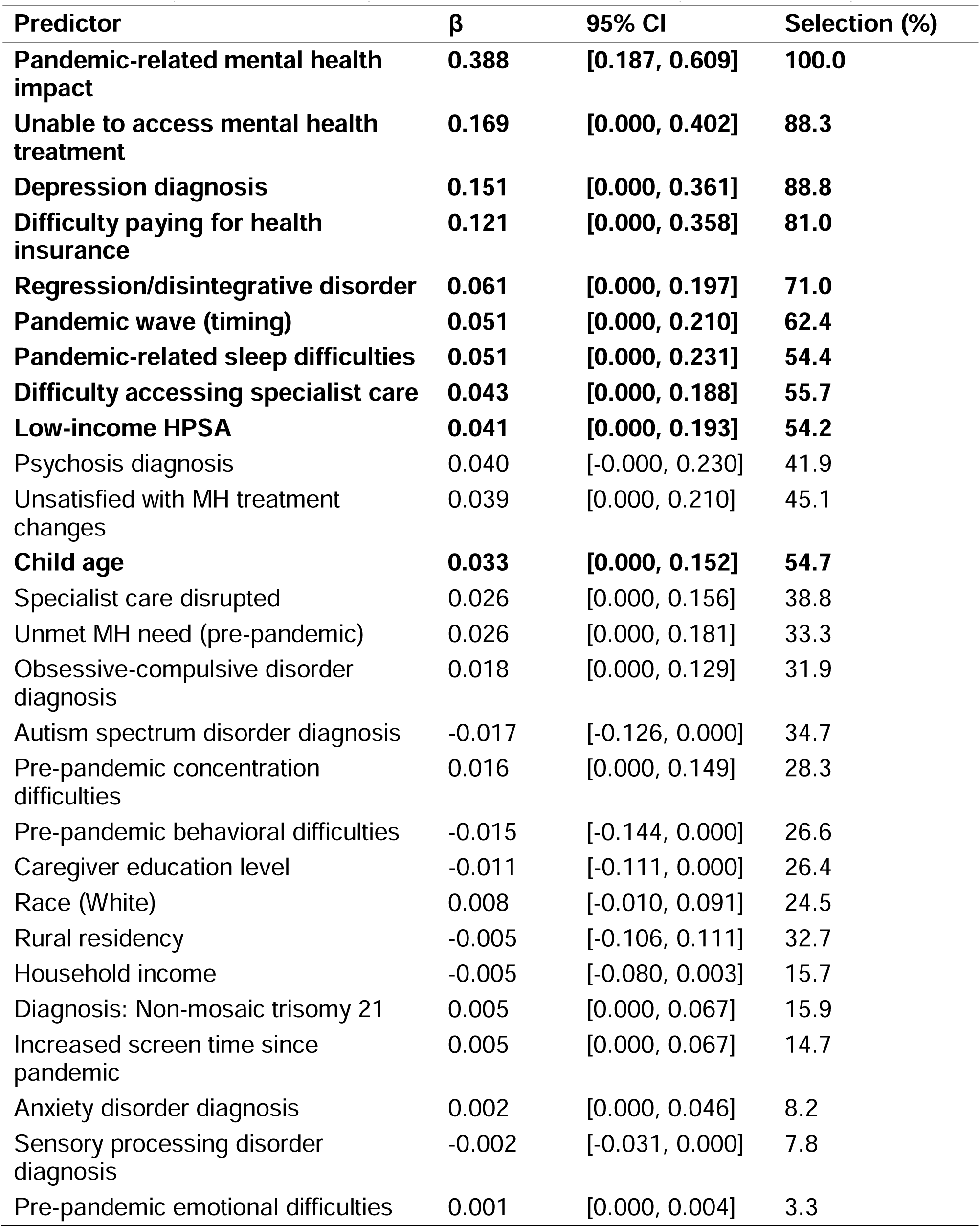
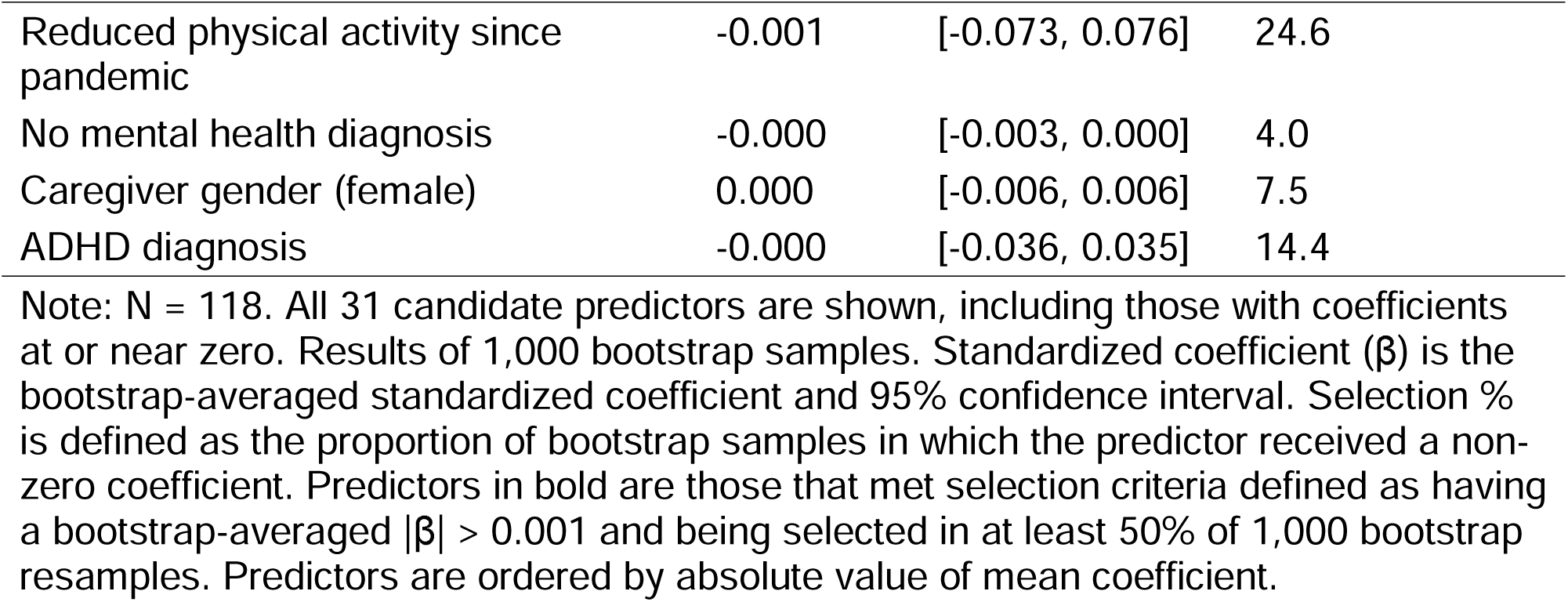
Elastic Net Regression Predicting Observed Functional Changes (NTG Changes)

| **Predictor** | **$\beta$** | **95% CI** | **Selection (%)** |
| --- | --- | --- | --- |
| **Pandemic-related mental health impact** | **0.388** | **[0.187, 0.609]** | **100.0** |
| **Unable to access mental health treatment** | **0.169** | **[0.000, 0.402]** | **88.3** |
| **Depression diagnosis** | **0.151** | **[0.000, 0.361]** | **88.8** |
| **Difficulty paying for health insurance** | **0.121** | **[0.000, 0.358]** | **81.0** |
| **Regression/disintegrative disorder** | **0.061** | **[0.000, 0.197]** | **71.0** |
| **Pandemic wave (timing)** | **0.051** | **[0.000, 0.210]** | **62.4** |
| **Pandemic-related sleep difficulties** | **0.051** | **[0.000, 0.231]** | **54.4** |
| **Difficulty accessing specialist care** | **0.043** | **[0.000, 0.188]** | **55.7** |
| **Low-income HPSA** | **0.041** | **[0.000, 0.193]** | **54.2** |
| Psychosis diagnosis | 0.040 | [-0.000, 0.230] | 41.9 |
| Unsatisfied with MH treatment changes | 0.039 | [0.000, 0.210] | 45.1 |
| **Child age** | **0.033** | **[0.000, 0.152]** | **54.7** |
| Specialist care disrupted | 0.026 | [0.000, 0.156] | 38.8 |
| Unmet MH need (pre-pandemic) | 0.026 | [0.000, 0.181] | 33.3 |
| Obsessive-compulsive disorder diagnosis | 0.018 | [0.000, 0.129] | 31.9 |
| Autism spectrum disorder diagnosis | -0.017 | [-0.126, 0.000] | 34.7 |
| Pre-pandemic concentration difficulties | 0.016 | [0.000, 0.149] | 28.3 |
| Pre-pandemic behavioral difficulties | -0.015 | [-0.144, 0.000] | 26.6 |
| Caregiver education level | -0.011 | [-0.111, 0.000] | 26.4 |
| Race (White) | 0.008 | [-0.010, 0.091] | 24.5 |
| Rural residency | -0.005 | [-0.106, 0.111] | 32.7 |
| Household income | -0.005 | [-0.080, 0.003] | 15.7 |
| Diagnosis: Non-mosaic trisomy 21 | 0.005 | [0.000, 0.067] | 15.9 |
| Increased screen time since pandemic | 0.005 | [0.000, 0.067] | 14.7 |
| Anxiety disorder diagnosis | 0.002 | [0.000, 0.046] | 8.2 |
| Sensory processing disorder diagnosis | -0.002 | [-0.031, 0.000] | 7.8 |
| Pre-pandemic emotional difficulties | 0.001 | [0.000, 0.004] | 3.3 |
| Reduced physical activity since pandemic | -0.001 | [-0.073, 0.076] | 24.6 |
| No mental health diagnosis | -0.000 | [-0.003, 0.000] | 4.0 |
| Caregiver gender (female) | 0.000 | [-0.006, 0.006] | 7.5 |
| ADHD diagnosis | -0.000 | [-0.036, 0.035] | 14.4 |

Note: N = 118. All 31 candidate predictors are shown, including those with coefficients at or near zero. Results of 1,000 bootstrap samples. Standardized coefficient ( $\beta$ ) is the bootstrap-averaged standardized coefficient and 95% confidence interval. Selection % is defined as the proportion of bootstrap samples in which the predictor received a non-zero coefficient. Predictors in bold are those that met selection criteria defined as having a bootstrap-averaged $|\beta| > 0.001$ and being selected in at least 50% of 1,000 bootstrap resamples. Predictors are ordered by absolute value of mean coefficient.

**Figure 3.**
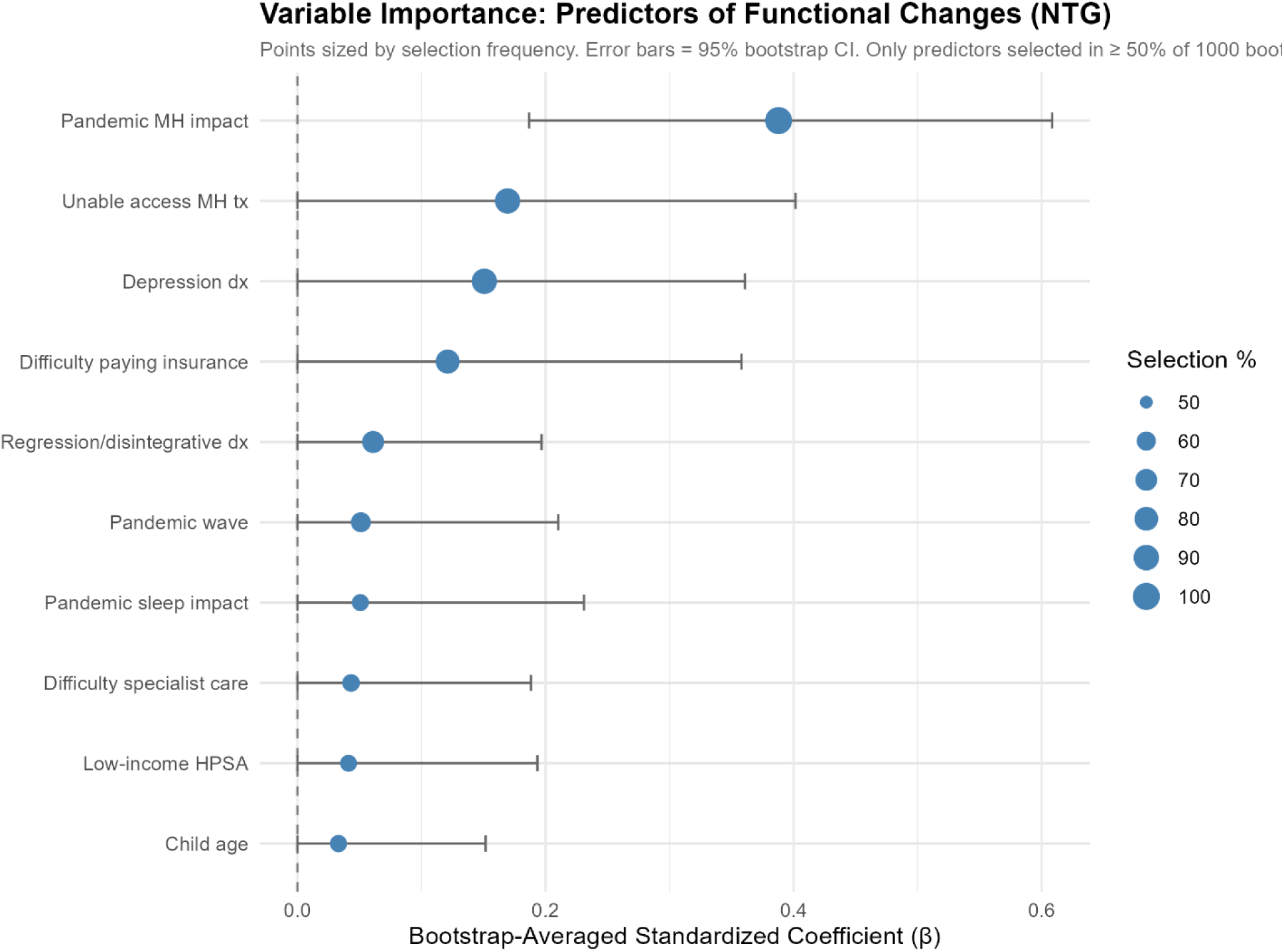
Variable importance plot for the 10 reliably retained predictors of functional changes. Points represent bootstrap-averaged standardized coefficients; horizontal bars represent 95% bootstrap percentile confidence intervals. Point size is proportional to selection frequency (the proportion of 1,000 bootstrap resamples in which each predictor received a non-zero coefficient).

The strongest and most consistently selected predictor included pandemic-related mental health impact (β = 0.388, 95% CI [0.187, 0.609]), which received a non-zero coefficient in 100% bootstrap samples. Four healthcare access barriers were also reliably retained: inability to access mental health treatment since the pandemic (β = 0.169, 95% CI [0.000, 0.402], 88.3%), difficulty paying for health insurance (β = 0.121, 95% CI [0.000, 0.358], 81.0%), difficulty accessing specialist care (β = 0.043, 95% CI [0.000, 0.188], 55.7%), and residing in low-income areas with health professional shortages (β = 0.041, 95% CI [0.000, 0.193], 54.2%). Pre-pandemic mental health diagnoses were also reliably selected, including a diagnosis of depression (β = 0.151, 95% CI [0.000, 0.361], 88.8%) and a regression or disintegrative disorder (β = 0.061, 95% CI [0.000, 0.197], 71.0%) were also among the more stably selected predictors. Additional retained predictors with modest contributions included pandemic wave timing (β = 0.051, 95% CI [0.000, 0.210], 62.4%), pandemic-related sleep impacts (β = 0.051, 95% CI [0.000, 0.231], 54.4%), and child age (β = 0.033, 95% CI [0.000, 0.152], 54.7%). The complete bootstrap-averaged coefficients for all 31 candidate predictors can be found in Table 2.

The model demonstrated moderate predictive performance, with a 3-fold cross-validated RMSE of 1.13, an R² of .35, and an MAE of 0.86. The RMSE indicates that, on average, the model’s predictions deviate slightly more than one unit from observed functional changes on the 0-5 outcome scale. The R² value indicates that approximately 35% of the variability in functional changes is explained by the model, highlighting a meaningful predictive capability but also suggesting that substantial unexplained variance remains. The MAE further indicates that typical prediction errors are less than one unit on the outcome scale.

## Discussion

This exploratory study examined the relationship between social determinants and mental health in the context of the COVID-19 pandemic, focusing on functional changes in individuals with Down syndrome. The study utilized gait, personality, friendliness, attention, and weight as indicators of these functional changes. The findings suggest a multifaceted relationship between functional changes, mental health, and healthcare access, aligning with a biopsychosocial model that emphasizes how biological vulnerabilities, psychological functioning, and social context collectively shape health outcomes in individuals with intellectual and developmental disabilities (IDD) (Bond et al., 2020).

Within this biopsychosocial framework, the strongest and most consistently selected predictor of functional changes was pandemic-related mental health impacts. A diagnosis of depression also emerged as one of the more stably selected predictors (selected in approximately 89% of bootstrap resamples), consistent with prior work documenting elevated rates of mood disorders in adults with Down syndrome (Mantry et al., 2007; Tassé et al., 2016) and reinforcing the importance of depression screening in this population. Although we cannot directly test causal associations between pre-existing mental health conditions and functional changes or whether the emergence of new functional changes caused worsening mental health symptoms, it is possible that the increased stress, social isolation, and disruptions to daily routines during the pandemic exacerbated pre-existing mental health conditions, leading to observable functional changes as has been observed in both the general population and individuals with intellectual disabilities (Hartley et al., 2022). Conversely, the appearance of new functional changes may be associated with heightened anxiety and depressive symptoms, creating a feedback loop that worsens clinical outcomes. Understanding this bidirectional dynamic requires longitudinal research to establish clearer temporal relationships. Other mental health diagnoses, such as regression, were also among the more stably selected predictors, further highlighting the complex biopsychosocial interplay between mental health and overall functioning in this population (Santoro et al., 2022; Sargado et al., 2022). Finally, combined sleep difficulties were also found to be associated with functional changes, indicating the broader effects of mental health and physiological challenges on functional changes (Capone, Aidikoff, Taylor, & Rykiel, 2013). Nevertheless, the directionality of these changes remains ambiguous, raising questions about the nature of interdependencies between functional capabilities and mental health concerns.

Our findings that mental health was closely associated with observable functional changes may offer valuable clues for caregivers and healthcare providers in identifying emerging mental health issues. Diagnostic overshadowing, a phenomenon where mental health symptoms are misattributed to Down syndrome or intellectual disability (ID), is a common challenge in this population (Mason & Scior, 2004). The findings suggest that functional changes could serve as an early signal for caregivers and providers to consider more thorough mental health assessments, as conventional tools may not always be effective in diagnosing mental health concerns in people with ID, particularly those with limited verbal skills (Spendelow, 2011). Future work should consider whether existing tools, such as the NTG-EDSD, can be used to screen for other mental health concerns, thereby increasing the efficiency of large-scale and phenotypically broad cohort studies.

In addition to informing improvements to mental health screening, our findings emphasize the intricate biopsychosocial relationship between mental health and overall functioning and highlight the importance of addressing mental health care for individuals with intellectual and developmental disabilities. There are many systemic barriers to effective treatments in this population, including limited access to care, social stigma, and a lack of mental health professionals trained to work with populations with intellectual and developmental disabilities (Werner & Stawski, 2012). Even within Down syndrome specialty clinics, there are considerable gaps in mental health services. A survey conducted by Santoro et al. (2021) across 14 Down syndrome specialty clinics found that while 50% of clinics identified having a psychologist as a “must-have”, only 7% had one available. This suggests that, even when individuals with Down syndrome can access physical healthcare, their mental health needs often go unmet. Further research should continue to develop adequate assessment tools for this population, which can inform to more appropriate, timely, and targeted interventions.

The study also highlights the role of healthcare accessibility in understanding the functional changes experienced by individuals with Down syndrome. Four healthcare access barriers were among the reliably retained predictors, including difficulty accessing mental health care during the pandemic, difficulty accessing specialist care, residing in a low-income or health professional shortage area, and difficulties affording health insurance. Even prior to the pandemic, accessing specialty care for people with Down syndrome was difficult, and many adults with Down syndrome continue to receive primary care from pediatricians well into adulthood, often because adult providers with relevant expertise are scarce (Jensen et al., 2021). Geospatial analyses have found that the majority of people with Down syndrome in the US do not have access to specialty clinics, with laws preventing services across state lines and rurality noted as significant barriers (Joslyn et al., 2020) After the onset of the pandemic, disruptions to disability support programs and healthcare services may have exacerbated these challenges, leaving caregivers with limited resources to address health and behavioral needs (Annaswamy et al., 2020; Hartley et al., 2022). The convergence of these access barriers reinforces the substantial biopsychosocial challenges faced by individuals with

Down syndrome in obtaining adequate healthcare. Interventions that are specifically tailored to the communication and cognitive needs of individuals with Down syndrome are needed (Vicari et al., 2013). A rapid shift to telehealth was a hallmark of the COVID-19 pandemic, presenting both new opportunities and additional challenges for many individuals with Down syndrome. Programs such as DSC2U have demonstrated the feasibility of remote multidisciplinary support for accessing Down syndrome–specific care (Majewski et al., 2021).

## Limitations and Implications for Future Research

This exploratory study highlights the need for further investigation into the complex relationships between social determinants, mental health, and functional changes in individuals with Down syndrome. Nevertheless, the present study is not without limitations. This study used a subscale of the NTG-EDSD, which was originally designed and validated for use a dementia screening tool, as a general measure of observable functional changes. The data for the present study was drawn from a rapid response surveillance study to understand the impacts of the pandemic on people with Down syndrome and their families, thus secondary measures were selected for theoretical relevance to health and well-being and for brevity to reduce participant burden. this study surveyed caregivers at a single time point over 2 years following the onset of the pandemic. Longitudinal studies could provide more clarity on the temporal dynamics of these relationships and help identify effective strategies for supporting this population during crises. Additionally, our findings suggest that future confirmatory research should examine whether improving access to mental health treatment during public health emergencies can mitigate functional changes in this population.

This study also relied on caregiver-reported data, which may introduce recall bias and subjective interpretations of functional changes. Moreover, while efforts were made to ensure diversity, the study’s reliance on online registries and Down syndrome-focused networks may limit its generalizability. The income distribution in our sample differs from the national distribution of U.S. families in 2020 (Board of Governors of the Federal Reserve System, 2021), which may limit the generalizability of our findings.

Specifically, our sample underrepresents lower-income households (16% earning <$50,000 compared to 45% nationally) and overrepresents middle-income households (66% earning $50,000–$99,999 compared to 25% nationally). While the proportion of high-income households (>$100,000) in our sample (30%) aligns closely with the national figure (29%), the underrepresentation of lower-income groups could introduce bias. Future research should aim to include a more diverse population, particularly those from racial and ethnic minority backgrounds and lower socioeconomic groups, to ensure a comprehensive understanding of the experiences of individuals with Down syndrome across varied contexts. Our analysis modeled only additive (main effect) contributions of predictors and did not test for interactions among them; future work with larger samples could examine whether the effects of access barriers or pandemic impacts differ by demographic or clinical subgroups.

## Conclusion

In conclusion, the findings of this study underscore the multifaceted challenges faced by individuals with Down syndrome during the COVID-19 pandemic, particularly regarding mental health and functional changes. Addressing these challenges requires not only tailored mental health interventions but also systemic changes to improve access to care and reduce barriers to services.

## Data Availability

All data produced in the present study are available upon reasonable request to the authors.

## Acknowledgments

The authors would like to thank the families who participated in this study for their willingness to share their experiences during the COVID-19 pandemic, as well as the many students who dedicated their time to this project.

## Ethics approval and consent to participate

This study was approved by the Institutional Review Board at Virginia Commonwealth University, and consent to participate was obtained from all parents or legal guardians of the participants.

## Author Contributions

R.B., N.W., L.A., C.J-C., A.B., G.C., and T.Y. were responsible for study design; R.B. acquired funding, performed data analysis and statistical reporting; E.J., C.W., and R.B. contributed substantially to writing the manuscript; R.B., A.D., and E.J. contributed substantially to data collection. All authors contributed substantially to the interpretation of results and edited and approved the manuscript.

